# Examining epidemiological models and economic analyses of Typhoid Conjugate Vaccine: A scoping review

**DOI:** 10.1101/2025.08.21.25334147

**Authors:** Prashant Mandaliya, Stacey Orangi, Jacob Kazungu, Dennis Waithaka, Angela Kairu, Amos Batiano, Felix Masiye, Obinna Onwujekwe, Edwine Barasa

## Abstract

Typhoid remains a preventable disease that primarily affects low- and middle-income countries. We conducted a scoping review to synthesize evidence on the epidemiological models and economic analyses of typhoid conjugate vaccine (TCV), focusing on the cost of illness of typhoid, the cost of vaccination, cost-effectiveness, and public health benefits of TCV.

A search was conducted across PubMed, Web of Science, International Health Technology Assessment, Scopus, and the National Health Service Economic Evaluation Database, following Arksey and O’Malleys’s framework. Screening and data extraction were managed using Covidence and an Excel-based template. Extracted data included study design, population, outcomes, model parameters, and cost-effectiveness data, converted to 2024, United States Dollars (USD) for comparability. Findings were summarized by key outcomes.

The economic burden of typhoid varied by region, care type, and demographics. Direct medical costs ranged from USD 1.3 to USD 1543, direct non-medical costs varied from USD 1.3 to USD 60.1, and indirect costs ranged from USD 14.1 to USD 794. Cost of vaccination per dose ranged from USD 2.80 - USD 5.40 (India), USD 0.87 - USD 1.01 (Malawi), and USD 2.99 (Zimbabwe), while the cost of delivering the vaccine ranged from USD 0.48- USD 0.99 (financial) and USD 1.11 - USD 1.87 (economic) per dose. Models predicted 2-94% case reductions and 0-100% mortality reductions. Economic evaluations reported ICERs of USD 124 to USD 53,773 per DALY averted and USD - 4,251 to USD 103,344 per QALY gained, with high-incidence settings often resulting in cost savings. Sensitive parameters were typhoid incidence rate, vaccine efficacy, and vaccination costs.

TCV introduction is influenced by disease burden, vaccination costs, and health outcomes, which vary by region. Cost-effectiveness depends on incidence, perspective, and vaccine strategy, emphasizing the need for context-specific evaluations. Targeted strategies, particularly in high-incidence and urban areas, are often cost-effective and sometimes cost-saving.

## Introduction

Typhoid fever remains an ongoing public health issue in low- and middle-income countries (LMICs), despite recent advances in preventive measures through vaccination. The disease is caused by *Salmonella enterica serovar Typhi* (*S. Typhi*). The primary route of transmission is by ingesting food or water that is contaminated with the pathogen [1]. Some individuals can become chronic carriers and may continue to shed the bacteria in their urine and feces for years, resulting in an increase in the rate of transmission [1].

Typhoid is endemic in Africa, Asia, Eastern Mediterranean, Latin America, Oceania, and the Western Pacific [2]. A commonly accepted threshold for defining high endemicity for typhoid or enteric fever is an incidence rate exceeding 100 cases per 100,000 persons per year [2]. Different modeling approaches have been adopted to estimate the overall burden of typhoid fever in each region. For instance, Antillón et al. estimated approximately 293 typhoid cases per 100,000 people in LMICs, with Central Africa predicted to have the highest incidence [3]. Another study estimated an annual incidence of 318 typhoid cases per 100,000 people in Africa [4]. Regarding the age distribution, a systematic review and meta-analysis of studies conducted in Africa and Asia reported that the proportion of typhoid fever cases was between 14-29% in children under five years old, 30-44% in those aged 5-9 years, and 28-52% in those aged 10-14 years [5]. For mortality, there were an estimated 110,000 deaths in 2019 were attributed to typhoid fever globally [6].

Initiation of treatment with the appropriate antibiotics leads to a case fatality rate for typhoid fever ranging from 1 to 4% [7]. However, this can increase to 10-20% if the condition is left untreated or if the inappropriate use of medications [7]. Although antibiotics have effectively treated typhoid fever, there has been a rise in multidrug-resistant strains posing a threat to disease management. Reports of *S. Typhi* strains resistant to fluoroquinolones and with increasing resistance to azithromycin are of concern because of the limited treatment options [8]. The combination of limited access to treatment in LMICs and antimicrobial resistance underscores the importance of preventive interventions such as vaccination, surveillance, behavioral changes, and those that improve sanitation [9].

In 2017, the World Health Organization’s (WHO) Strategic Advisory Group of Experts (SAGE) on Immunization recommended the inclusion of TCV in the routine immunization programs of endemic countries [10]. The three types of WHO-approved typhoid vaccines are typhoid conjugate vaccine (TCV), unconjugated Vi polysaccharide (ViPS), and live attenuated Ty21a [11]. Although ViPS and Ty21a vaccines have been available since the early 2000s, there was a low uptake due to factors like lower efficacy, the requirement for multiple doses, and being contraindicated for use in children under two years of age [11,12]. In contrast, the newer TCV offers several advantages over its predecessors, including an efficacy greater than 80%, protection lasting over five years, a single-dose regimen, and being recommended for use in children as young as six months. These benefits have made TCV a strong candidate for inclusion in routine immunization programs in LMICs [11,13–17].

However, the available literature on the economic evaluation of TCV is still limited. Economic evaluations of vaccines involve a range of analyses that assess both economic and health-related outputs, which provide insights that can be used in public health policy decision-making. A comprehensive evaluation typically includes assessments of cost-of-illness (COI), which quantifies the total economic burden of a disease by estimating direct and indirect costs [18]. Studies on the costs of vaccine procurement and delivery examine the financial resources necessary for implementing vaccination programs [19]. Furthermore, modeling studies also play a vital role in evaluating vaccination strategies through simulations of potential outcomes over time and under different conditions, offering insights into their long-term effects [19]. Finally, cost-effectiveness analysis (CEA) systematically compares the costs and health outcomes (effectiveness) of different interventions, which can be used to determine the economic value of the intervention, which can help guide resource allocation [19]. Together, these components provide a comprehensive understanding of the economic and societal value vaccines offer, helping immunization programs to achieve the most significant public health impact possible.

A review by Luthra et al. in 2019 offered initial insights into the cost of illness of typhoid, the cost of vaccine delivery, cost-effectiveness, and demand forecast of the different types of typhoid vaccines [19]. However, since the WHO SAGE recommendation and prequalification of TCV in 2018, along with the Global Alliance for Vaccines and Immunization (GAVI) subsequent commitment to support its introduction, there has been an increase in country-level interest in economic evaluations assessing the cost-effectiveness, health impact, and potential cost savings associated with the use of TCV [19–21]. Despite this growing body of research, no recent scoping review has been conducted to systematically organize and synthesize the evidence. A scoping review would help fill the gap by mapping the current research landscape on the economic implications of TCV, identifying knowledge gaps, and highlighting areas that require further study. This review type is well-suited, as it allows for a broad exploration and systematic organization of the literature on TCV’s economic evaluation conducted over a relatively short period. Unlike systematic reviews or meta-analyses, which primarily assess quality or effectiveness, a scoping review identifies, organizes, and maps the breadth and nature of research in a field [22].

This review aimed to fill this gap by organizing and analyzing evidence on the economic evaluation of typhoid conjugate vaccine (TCV). It synthesized findings from a range of epidemiological models and economic analyses, covering different modeling approaches, vaccination strategies, costs related to both illness and immunization, public health outcomes, and cost-effectiveness. The primary focus of the review was on TCV given its benefits over other Typhoid vaccines that are available. This review can be used by both researchers and policymakers to get a better understanding of the economic value of TCV and use the evidence for decision-making in typhoid-endemic regions and aid in TCV rollout at a population level.

## Materials and Methods

This scoping review adhered to the methodological framework outlined by Arksey and O’Malley [23], which organized the process into five stages: [1] identification of the research question, [2] identification of relevant studies, [3] selection of eligible studies, [4] data extraction, and [5] collation, summarization, and reporting of the results. Additionally, the review followed the Preferred Reporting Items for Systematic Reviews and Meta-Analyses extension for scoping reviews (PRISMA-ScR) [24] as the reporting guideline to ensure transparency throughout the review process.

The review protocol was uploaded to the Open Science Framework (OSF): https://doi.org/10.17605/OSF.IO/WXDEU

### 1. Identification of the Research Question

This review addressed the research question: What evidence exists regarding the economic and public health impact of implementing the typhoid conjugate vaccine in a population?

By identifying studies evaluating these impacts through various methods, the review particularly focused on those reporting relevant outcomes such as cases averted, deaths averted, costs, incidence or transmission reduction, life-years gained, or an incremental cost-effectiveness ratio (ICER). This comprehensive research question guided the systematic search and selection of relevant studies.

### 2. Identification of Relevant Studies

An initial search was conducted on PubMed using key terms such as “Costs,” “Cost-effectiveness,” “Model,” “Epidemiological Model,” and “Typhoid conjugate vaccine.” After reviewing the titles and abstracts of the search results, additional relevant keywords were incorporated to refine the search strategy, as summarized in Table 1. The detailed strategy is found in S1 Appendix.

**Table 1:**
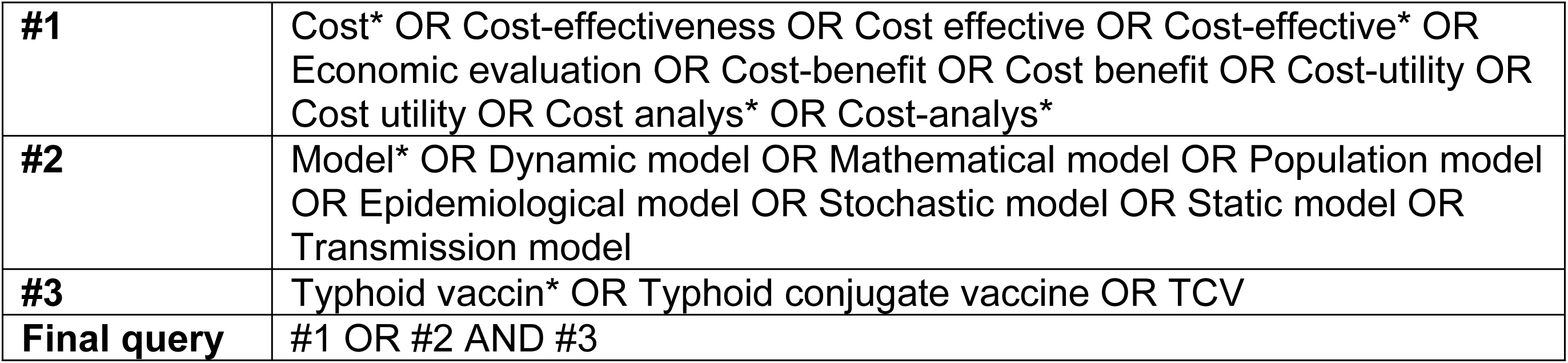
Search strategy.

|  |  |
| --- | --- |
| **#1** | Cost* OR Cost-effectiveness OR Cost effective OR Cost-effective* OR Economic evaluation OR Cost-benefit OR Cost benefit OR Cost-utility OR Cost utility OR Cost analys* OR Cost-analys* |
| **#2** | Model* OR Dynamic model OR Mathematical model OR Population model OR Epidemiological model OR Stochastic model OR Static model OR Transmission model |
| **#3** | Typhoid vaccin* OR Typhoid conjugate vaccine OR TCV |
| **Final query** | #1 OR #2 AND #3 |

The refined search strategy was then applied by two authors across several databases, including PubMed, Web of Science, International Health Technology Assessment (HTA), Scopus, and the National Health Service Economic Evaluation Database (NHS EED). Reference lists of included studies were screened, and Google Scholar was used to identify additional literature. The final database search was done on January 20, 2025.

### 3. Selection of Eligible Studies

The results from the database searches were imported into Covidence [25] for duplicate removal, screening of titles and abstracts, and full-text review. Titles and abstracts were independently reviewed by two reviewers using the inclusion and exclusion criteria in Table 2. Screening happened in two stages. First, reviewers assessed the titles and abstracts to flag studies that might be relevant. If a title seemed relevant but the abstract didn’t provide enough detail, the study was kept for full-text review to avoid missing important studies. Next, full texts were reviewed to decide whether each study met the criteria. For any studies that were excluded at this stage, the reason was recorded to keep the process transparent.

**Table 2:** Inclusion criteria.

| Criteria | Inclusion | Exclusion |
| --- | --- | --- |
| **Language** | English | Non-English |
| **Publication Type** | Peer-reviewed articles, full text available | Grey literature, full text unavailable |
| **Model requirement** | Model parameters must directly include TCV | Models lacking TCV-specific parameters |
| **Study Methodology** | Quantitative studies with empirical data | Qualitative studies, Opinion pieces, editorials, literature reviews, books |
| **Outcomes** | -Cost of illness of typhoid<br>- Cost of vaccine delivery/vaccination for TCV<br>- Public health impact of TCV<br>- Cost-effectiveness of TCV | Studies without these specific outcomes |

The study selection followed the PRISMA-ScR guidelines [24] (S2 Appendix), and the process is outlined in a PRISMA flow diagram (Fig 1) to support transparency and reproducibility. If the two reviewers disagreed at any point, a third reviewer was consulted to help resolve the differences and reach a consensus.

**Fig 1.**
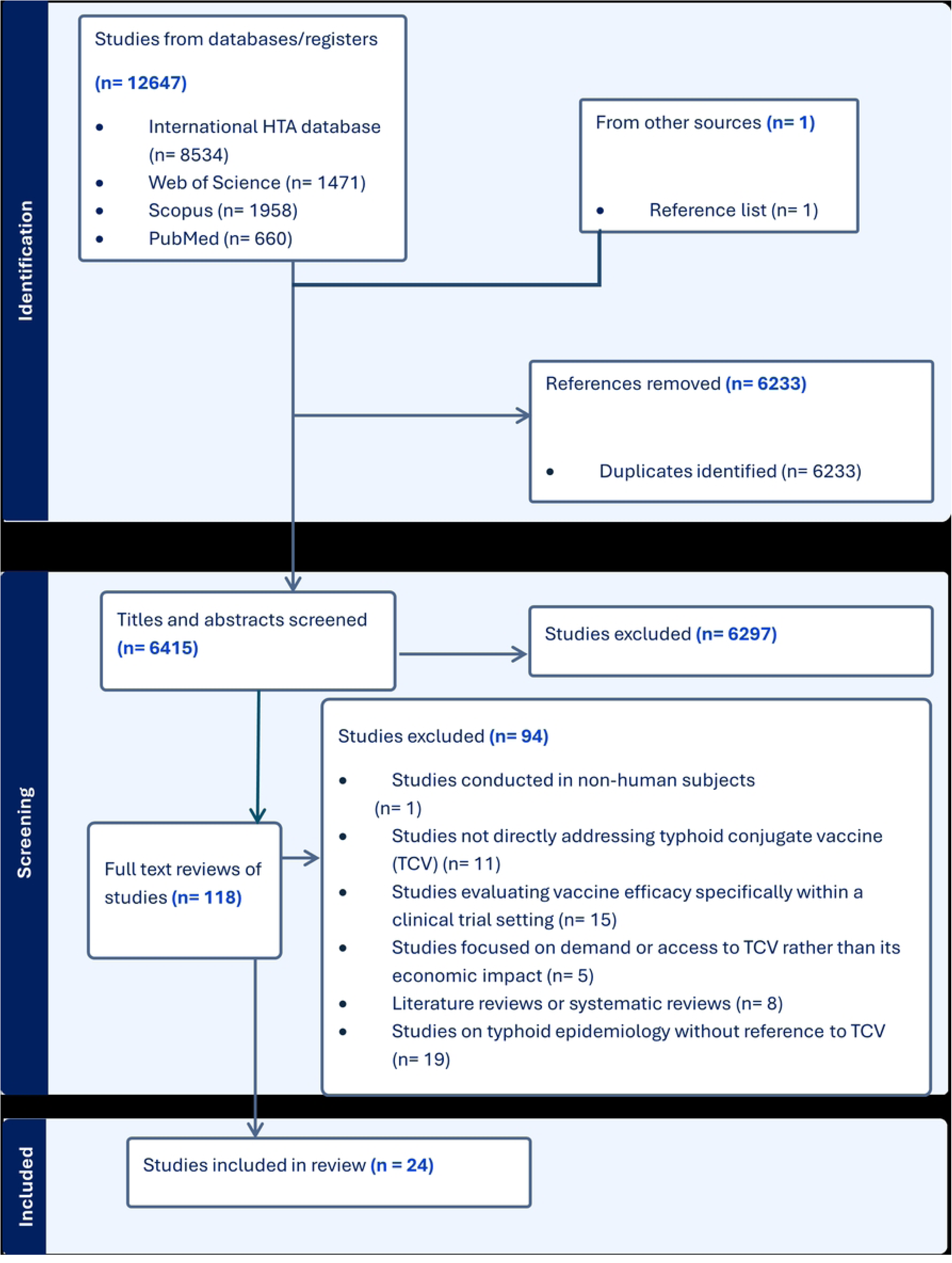
Economic and public health impact PRISMA flow diagram. Flow diagram illustrating the number of records identified, screened, included, and excluded during the study.

### 4. Data extraction

Data from the 24 included studies were extracted using a structured method to ensure accuracy and completeness. A standardized Microsoft Excel template (S3 Appendix) was used to record key details from each study, including the title, lead author, year of publication, vaccination strategies, model type, time horizon, perspective, cycle length, and any parameter stratification used in the epidemiological model. The template also captured reported values on the cost of illness, vaccination costs, public health outcomes, ICERs, and other major findings.

### 5. Collation, Summarization, and Reporting of Results

The analysis included a descriptive summary of the quantitative data, sorted into four sub-groups: i) cost of illness, ii) cost of vaccination, iii) public health impact, and iv) economic evaluation. Results for each sub-group were organized into separate tables based on the primary outcomes. This consistent structure made it easier to identify trends and draw conclusions about the economic and public health impact of TCV for decision-makers.

#### Cost Adjustment Over Time

To allow comparison across studies, all monetary values were adjusted to 2024 USD using Consumer Price Index (CPI) data [26]. For the study [27] that reported values in Indian rupees (INR), we first converted the reported INR value to 2015 USD using an exchange rate of 70.4 [28], and then adjusted it to 2024 USD. For the study that reported monetary values in 2015 international dollars [29], we directly adjusted these to 2024 USD using CPI. This approach assumed that 1 international dollar is equivalent to 1 USD, which is consistent with The World Bank’s definition [30].

#### Quality assessment

The economic evaluations were assessed using the Drummond 10-point checklist [31] (S4 Appendix), a tool used for evaluating economic evaluations of health interventions. The process involved two reviewers independently scoring each study by awarding one point for each criterion met. Based on Doran’s classification system [32], studies were then rated as low (1–3), moderate (4–7), or high quality (8–10 ), based on the score they received.

For studies focusing on the public health impact of TCV, a tool that assessed infectious disease models was considered [33]. However, this tool mainly focused on aspects of the software used and data availability, which made it unsuitable for our review.

No standardized quality assessment tool was identified for cost of illness or cost of vaccination studies. We therefore assumed that peer-reviewed publications in these categories met acceptable quality standards. This approach helped maintain consistency across the different types of studies included in this review.

All included economic evaluations were rated as high quality, with full details shown in Table 3.

**Table 3:** Quality assessment of included cost-effectiveness analyses using the Drummond checklist [31] and Doran [32] scoring system.

|  | Antillo<br>n et al | Bilcke<br>et al | Burrow<br>s et al | Chauha<br>n et al | Lo<br>NC et<br>al | Phillip<br>s et al | Ryckma<br>n et al | Soukavon<br>g et al | Weyan<br>t et al |
| --- | --- | --- | --- | --- | --- | --- | --- | --- | --- |
| 1. Was a well-defined question posed in answerable form? | 1 | 1 | 1 | 1 | 1 | 1 | 1 | 1 | 1 |
| 2. Was a comprehensive description of the competing alternatives given? | 1 | 1 | 1 | 1 | 1 | 1 | 1 | 1 | 1 |
| 3. Was the effectiveness of the programme established? | 1 | 1 | 1 | 1 | 1 | 1 | 1 | 1 | 1 |
| 4. Were all the important and relevant costs and consequences for each alternative identified? | 1 | 1 | 1 | 1 | 1 | 1 | 1 | 1 | 1 |
| 5. Were costs and consequences measured accurately in appropriate physical units? | 1 | 1 | 1 | 1 | 1 | 1 | 1 | 1 | 1 |
| 6. Were costs and consequences valued credibly? | 1 | 1 | 1 | 1 | 1 | 1 | 1 | 1 | 1 |
| 7. Were costs and consequences adjusted for differential timing? | 1 | 1 | 1 | 1 | 1 | 1 | 1 | 1 | 1 |
| 8. Was an incremental analysis of costs and consequences of alternatives performed? | 1 | 1 | 1 | 1 | 1 | 1 | 1 | 1 | 1 |
| 9. Was allowance made for uncertainty in the estimates of costs and consequences? | 1 | 1 | 1 | 1 | 1 | 1 | 1 | 0 | 1 |
| 10. Did the presentation and discussion of study results include all issues of concern to users? | 1 | 1 | 1 | 1 | 1 | 1 | 1 | 1 | 1 |
| **Total** | 10 | 10 | 10 | 10 | 10 | 10 | 10 | 9 | 10 |
| **Quality level** | GOOD | GOOD | GOOD | GOOD | GOOD | GOOD | GOOD | GOOD | GOOD |
| **KEY:** The checklist can be used to rate the quality of an economic evaluation on a scale of 1–10, with 1–3 points indicating poor quality, 4–7 points indicating average quality, and 8–10 points indicating good quality. |  |  |  |  |  |  |  |  |  |

## Results

### General characteristics of the included studies

We included 24 studies published between 1998 and 2024, with 6 studies being multi-country analyses. Across the 24 studies, the number of distinct countries represented per region were: Sub-Saharan Africa (36 countries), South Asia (8 countries), East Asia and the Pacific (13 countries), Europe and Central Asia (8 countries), Latin America and the Caribbean (6 countries), and the Middle East and North Africa (4 countries). Two studies looked at LMICs where typhoid is endemic [34,35].

Of the 24 studies, eight assessed the COI for typhoid [36–43], four estimated the cost of TCV vaccination [44–47], one estimated the cost of TCV delivery [48], two modeled public health impact [34,49] and nine conducted economic evaluations [20,27,29,35,50–54].

Among the eight COI studies, seven adopted a societal or patient perspective [37,39–43], while one combined the patient and provider perspective [38]. In terms of the study population, five studies included patients of all ages [36–39,41], one focused on infants older than 2 months [43], another one included patients over 6 months [42], and one used different age groups for different countries [40].

Four studies estimated the cost of vaccination, and one study estimated the cost of delivery for TCV [44–48]. These studies employed various delivery strategies, including routine, campaign, and mixed. In India, campaigns targeted children aged 9 months to 14–15 years [44,55]; in Malawi, routine immunization (children >9 months) was combined with campaigns for children aged 9 months to <15 years [45,48]; and in Zimbabwe, campaigns alone targeted children aged from 6 months to 45 years [46].

The two public health impact studies assessed TCV strategies over a 10-year horizon using dynamic transmission models [34,49]. This was the same time horizon used by seven of the nine economic evaluations [20,29,35,50,51,53,54], while of the remaining two, one employed a 15-year horizon [27], and one adopted a life-time horizon [52]. The information was summarised and presented in Table 4.

**Table 4:**
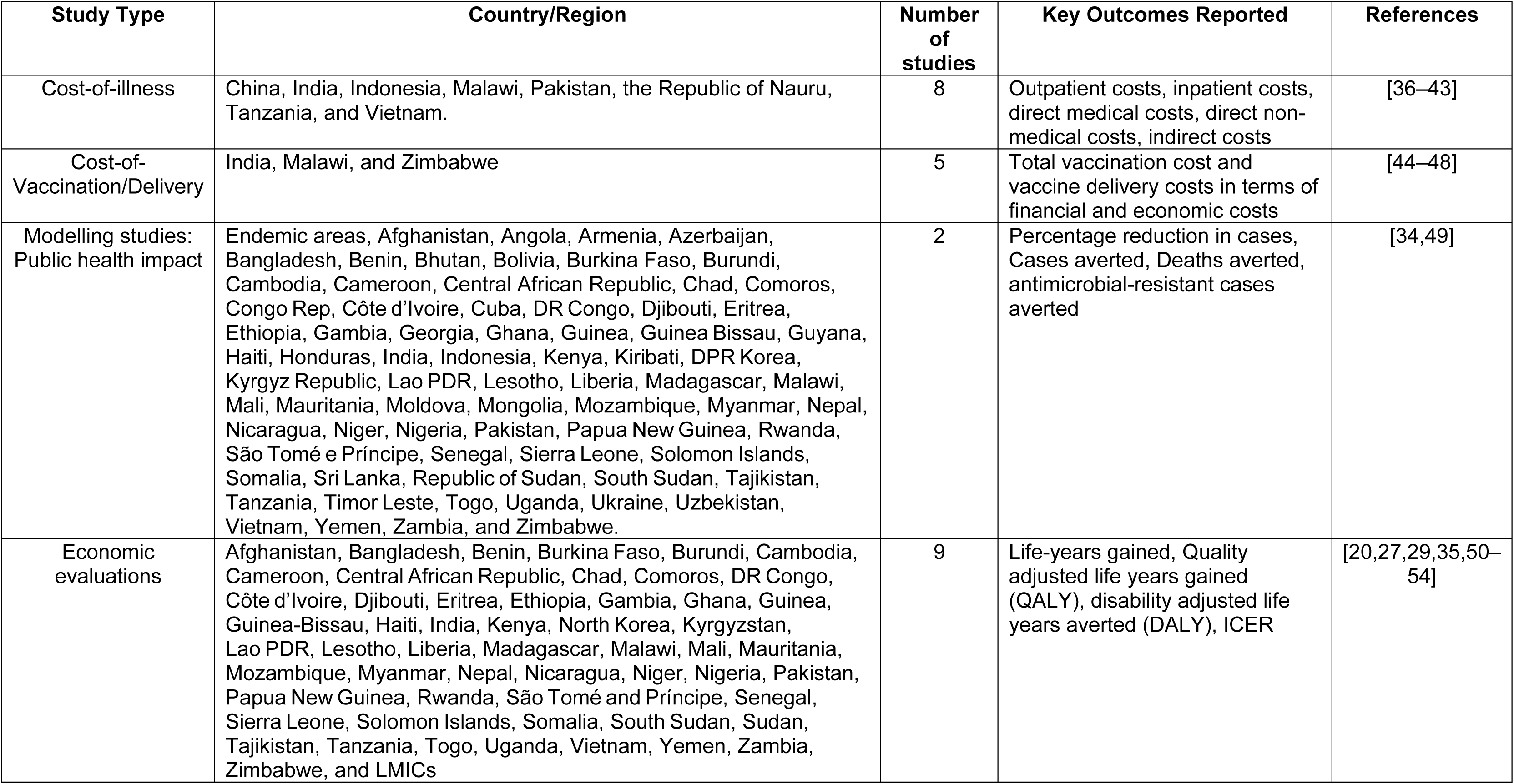
Summary of included studies.

S5 Table (Supporting information) provides a comprehensive overview of all extracted studies, capturing detailed unadjusted cost values from each study’s findings before similar findings were grouped.

### Cost of illness

Eight of the included studies [36–43] examined the economic burden of typhoid fever. The studies were summarized by location, perspective, population, cost parameters, and key findings. The variation in outcomes between the studies depended on factors such as the type of costs included (direct medical, non-medical, and/or indirect), region, patient demographics, and healthcare setting.

The reported total outpatient costs ranged from USD 5.4 to an estimated USD 300 [37,38,40,41], with significant differences across regions. In Sub-Saharan Africa, the reported outpatient cost was USD 49.5 [37]; in South Asia, from USD 5.4 to USD 61 [38,40], and in East Asia and the Pacific, from USD 54.5 to USD 300 [40,41]. Inpatient treatment costs were consistently higher than outpatient costs, ranging from USD 79 to USD 1750 [36–43]. Regional comparisons showed inpatient costs of USD 193-

437.2 in Sub-Saharan Africa [37,43], USD 79 - 636.7 in South Asia [36,38,40,42], and USD 253 - 1750 in East Asia and the Pacific [39–41]. Inter-regional differences in East Asia and the Pacific were highlighted by Poulos et al. [40], where the inpatient cost in Vietnam was USD 253 compared to USD 695 in Indonesia.

The direct medical costs varied by age and type of care (outpatient/ inpatient). Outpatient direct medical costs ranged from USD 1.3 to USD 157.4 [37,38,40]. When comparing age groups, children under 15 years had reported costs of USD 1.3 [37], while adults had higher direct medical costs of USD 1.4 – 157.4 [37,38,40]. The direct medical costs for inpatients ranged from USD 2.5 to USD 1543 [36–39,43], with estimates for children of USD 2.9 [37], and those for adults ranged from USD 2.5 to USD 1543 [36,38,39,43].

Direct non-medical costs, mainly transport, were higher for inpatients (USD 22.5 to USD 60.1) [36–38,43] than outpatients (USD 1.3 to USD 5.7) [37,38] and were more substantial in regions with limited access to healthcare.

The economic burden of typhoid fever extends beyond direct medical costs, with indirect costs (lost productivity) being significant, particularly for hospitalized patients. For outpatients, the indirect costs ranged from USD 14.1 to USD 34.2, as reported by one study [37]. In contrast, indirect costs for inpatients showed greater variation, ranging from USD 17.7 to USD 794 [36–39,42,43]. In some studies, inpatient indirect costs even exceeded the direct medical costs [37–39,43].

Complications of typhoid fever further increase the financial burden. Three studies [38,41,42] explored the inpatient costs associated with treating these complications. Punjabi [41], estimated costs of treating severe cases to exceed USD 490. Mejia et al. [38] reported the cost of treating intestinal perforation, which is a life-threatening complication, with estimated treatment costs of USD 636. Finally, Gupta et al. [42] reported the cost of managing sequelae, death, or referred cases as USD 139. The detailed findings are presented in Table 5.

**Table 5:**
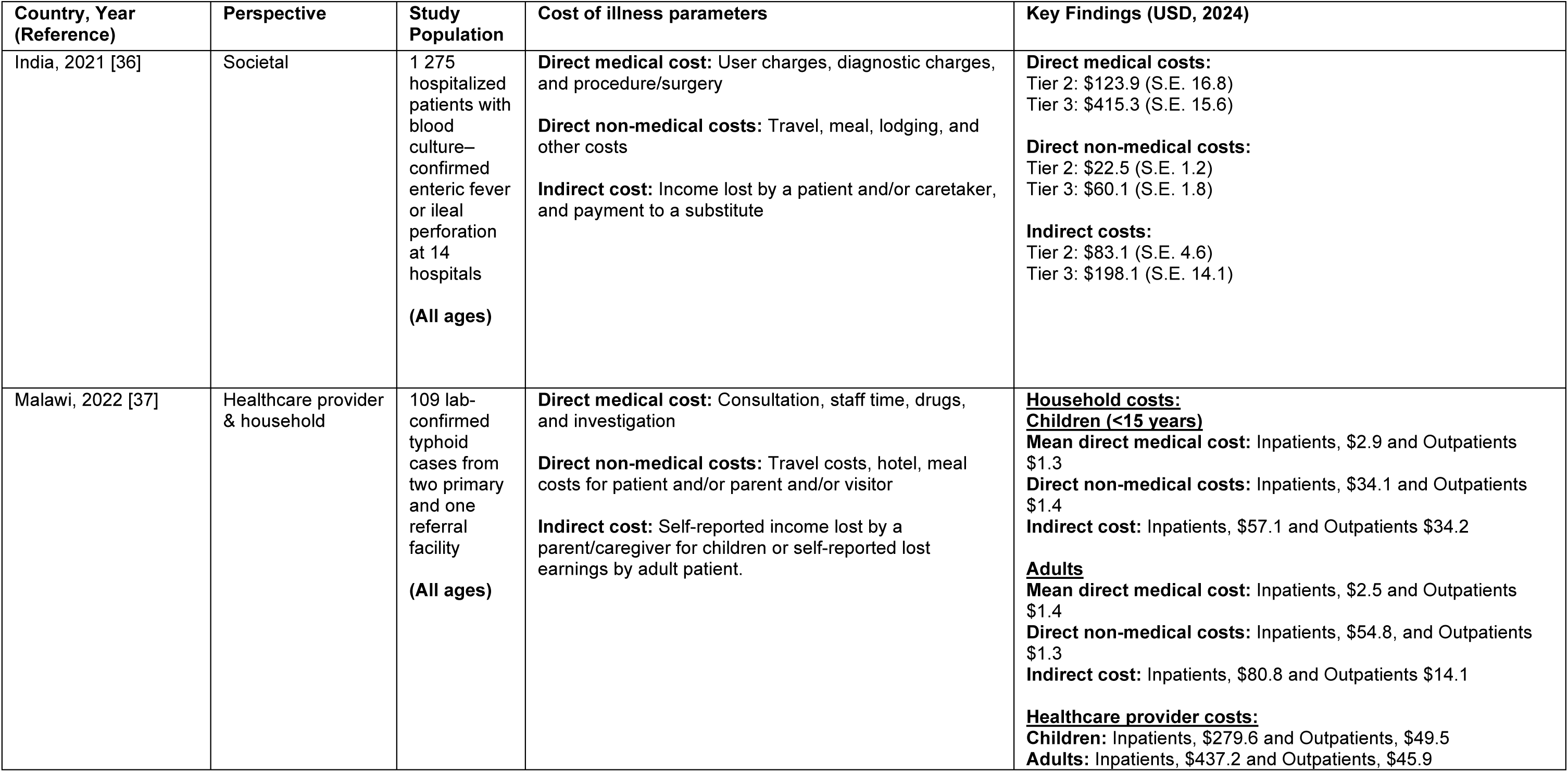
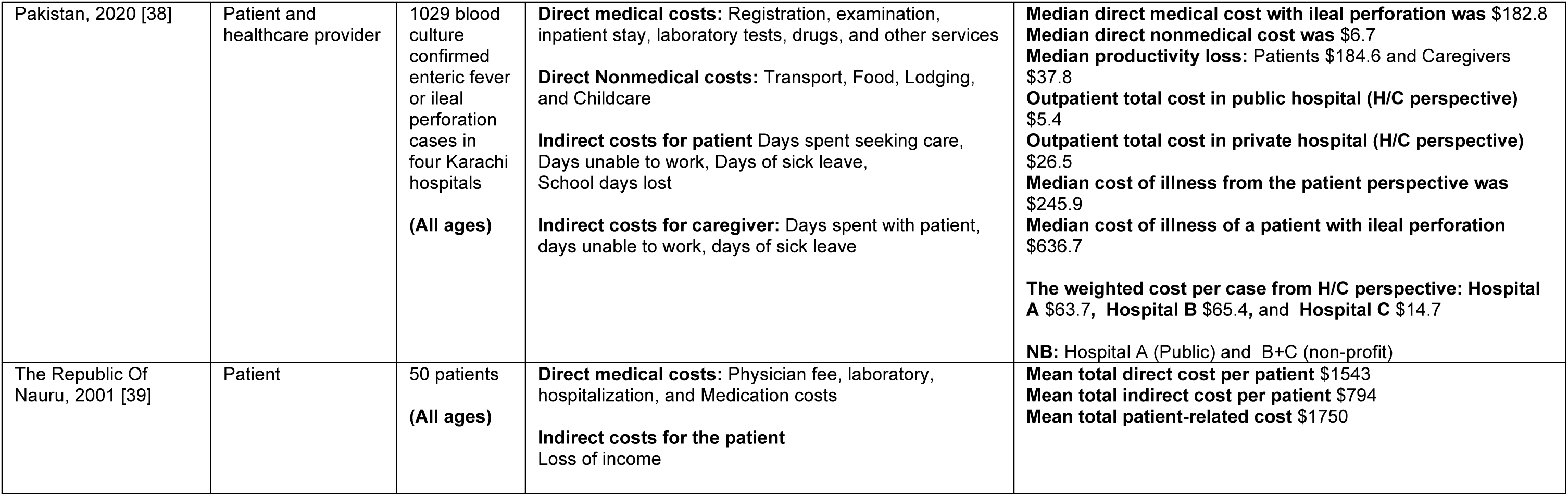
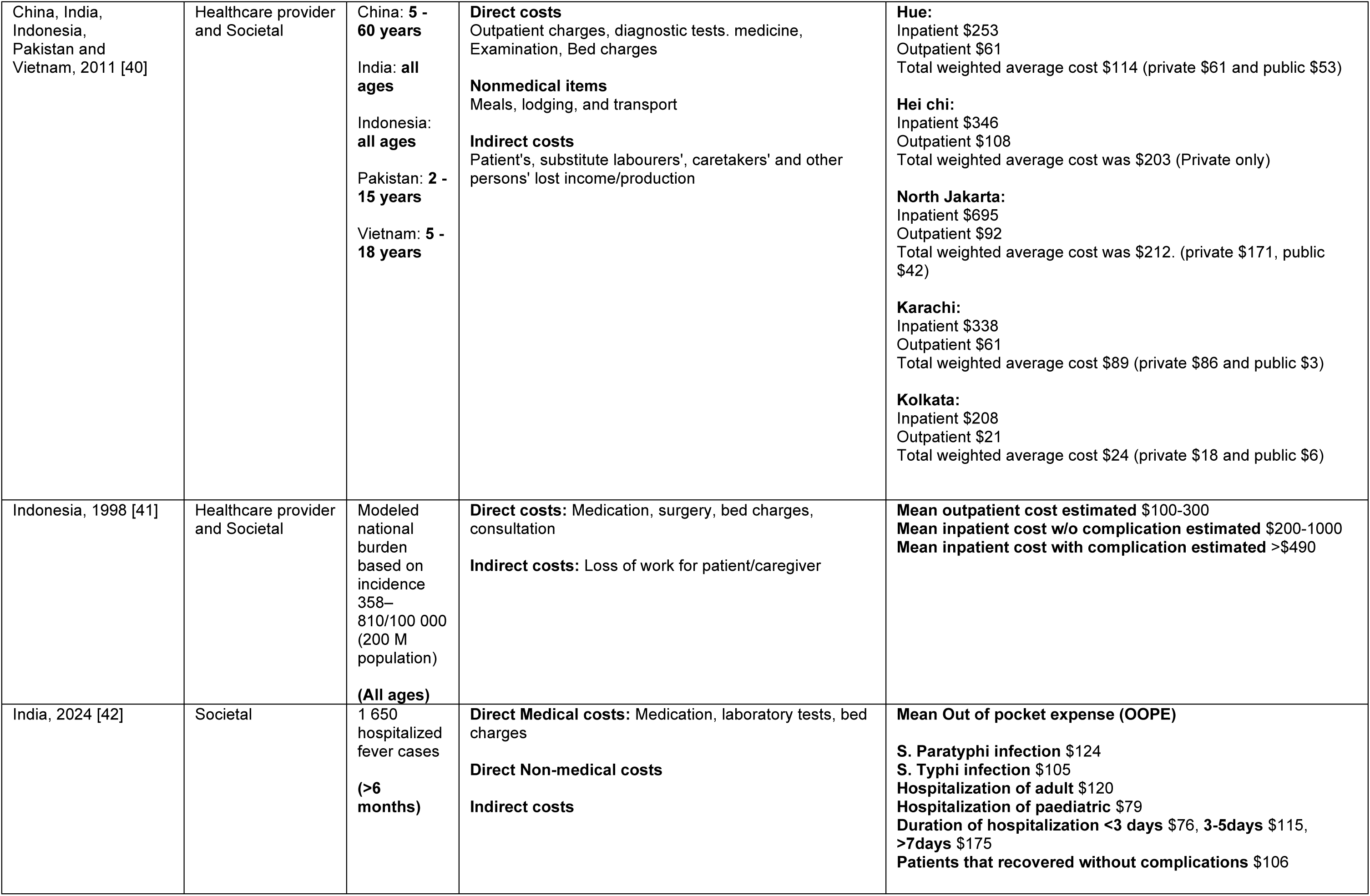
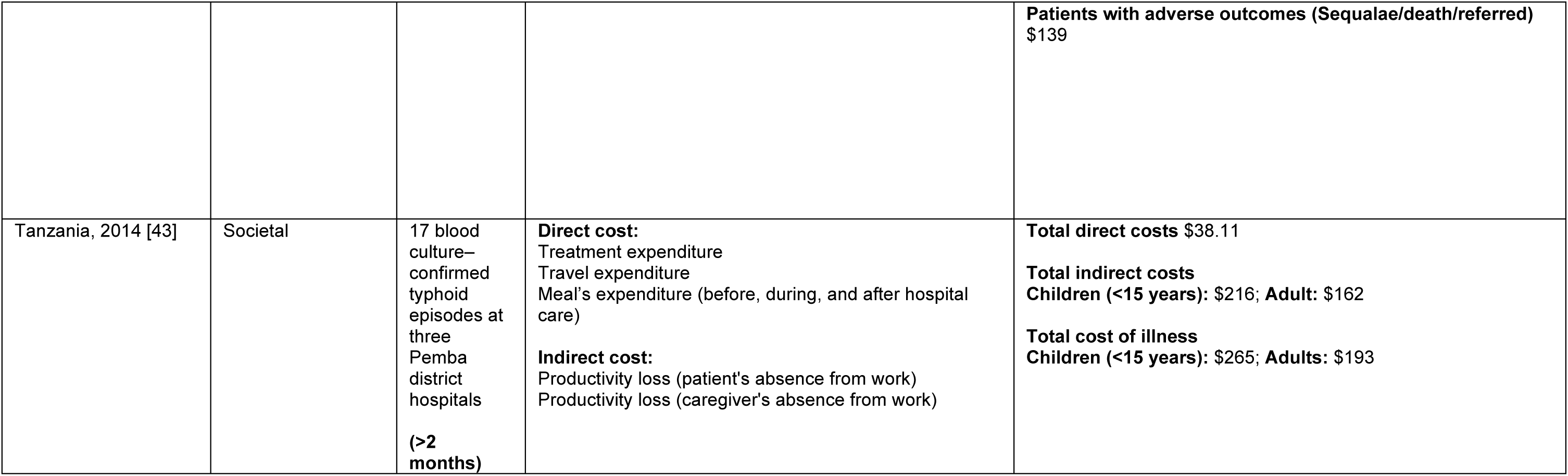
Summary of Cost of illness of typhoid studies.

| Country, Year (Reference) | Perspective | Study Population | Cost of illness parameters | Key Findings (USD, 2024) |
| --- | --- | --- | --- | --- |
| India, 2021 [36] | Societal | 1 275 hospitalized patients with blood culture–confirmed enteric fever or ileal perforation at 14 hospitals<br>(All ages) | **Direct medical cost:** User charges, diagnostic charges, and procedure/surgery<br>**Direct non-medical costs:** Travel, meal, lodging, and other costs<br>**Indirect cost:** Income lost by a patient and/or caretaker, and payment to a substitute | **Direct medical costs:**<br>Tier 2: \$123.9 (S.E. 16.8)<br>Tier 3: \$415.3 (S.E. 15.6)<br>**Direct non-medical costs:**<br>Tier 2: \$22.5 (S.E. 1.2)<br>Tier 3: \$60.1 (S.E. 1.8)<br>**Indirect costs:**<br>Tier 2: \$83.1 (S.E. 4.6)<br>Tier 3: \$198.1 (S.E. 14.1) |
| Malawi, 2022 [37] | Healthcare provider & household | 109 lab-confirmed typhoid cases from two primary and one referral facility<br>(All ages) | **Direct medical cost:** Consultation, staff time, drugs, and investigation<br>**Direct non-medical costs:** Travel costs, hotel, meal costs for patient and/or parent and/or visitor<br>**Indirect cost:** Self-reported income lost by a parent/caregiver for children or self-reported lost earnings by adult patient. | **Household costs:**<br>**Children (<15 years)**<br>**Mean direct medical cost:** Inpatients, \$2.9 and Outpatients \$1.3<br>**Direct non-medical costs:** Inpatients, \$34.1 and Outpatients \$1.4<br>**Indirect cost:** Inpatients, \$57.1 and Outpatients \$34.2<br>**Adults**<br>**Mean direct medical cost:** Inpatients, \$2.5 and Outpatients \$1.4<br>**Direct non-medical costs:** Inpatients, \$54.8, and Outpatients \$1.3<br>**Indirect cost:** Inpatients, \$80.8 and Outpatients \$14.1<br>**Healthcare provider costs:**<br>**Children:** Inpatients, \$279.6 and Outpatients, \$49.5<br>**Adults:** Inpatients, \$437.2 and Outpatients, \$45.9 |
| Pakistan, 2020 [38] | Patient and healthcare provider | 1029 blood culture confirmed enteric fever or ileal perforation cases in four Karachi hospitals<br>**(All ages)** | **Direct medical costs:** Registration, examination, inpatient stay, laboratory tests, drugs, and other services<br>**Direct Nonmedical costs:** Transport, Food, Lodging, and Childcare<br>**Indirect costs for patient** Days spent seeking care, Days unable to work, Days of sick leave, School days lost<br>**Indirect costs for caregiver:** Days spent with patient, days unable to work, days of sick leave | **Median direct medical cost with ileal perforation was \$182.8**<br>**Median direct nonmedical cost was \$6.7**<br>**Median productivity loss:** Patients \$184.6 and Caregivers \$37.8<br>**Outpatient total cost in public hospital (H/C perspective) \$5.4**<br>**Outpatient total cost in private hospital (H/C perspective) \$26.5**<br>**Median cost of illness from the patient perspective was \$245.9**<br>**Median cost of illness of a patient with ileal perforation \$636.7**<br>**The weighted cost per case from H/C perspective: Hospital A \$63.7, Hospital B \$65.4, and Hospital C \$14.7**<br>**NB:** Hospital A (Public) and B+C (non-profit) |
| The Republic Of Nauru, 2001 [39] | Patient | 50 patients<br>**(All ages)** | **Direct medical costs:** Physician fee, laboratory, hospitalization, and Medication costs<br>**Indirect costs for the patient**<br>Loss of income | **Mean total direct cost per patient \$1543**<br>**Mean total indirect cost per patient \$794**<br>**Mean total patient-related cost \$1750** |
| China, India, Indonesia, Pakistan and Vietnam, 2011 [40] | Healthcare provider and Societal | China: **5 - 60 years**<br>India: **all ages**<br>Indonesia: **all ages**<br>Pakistan: **2 - 15 years**<br>Vietnam: **5 - 18 years** | **Direct costs**<br>Outpatient charges, diagnostic tests. medicine, Examination, Bed charges<br>**Nonmedical items**<br>Meals, lodging, and transport<br>**Indirect costs**<br>Patient's, substitute labourers', caretakers' and other persons' lost income/production | **Hue:**<br>Inpatient \$253<br>Outpatient \$61<br>Total weighted average cost \$114 (private \$61 and public \$53)<br>**Hei chi:**<br>Inpatient \$346<br>Outpatient \$108<br>Total weighted average cost was \$203 (Private only)<br>**North Jakarta:**<br>Inpatient \$695<br>Outpatient \$92<br>Total weighted average cost was \$212. (private \$171, public \$42)<br>**Karachi:**<br>Inpatient \$338<br>Outpatient \$61<br>Total weighted average cost \$89 (private \$86 and public \$3)<br>**Kolkata:**<br>Inpatient \$208<br>Outpatient \$21<br>Total weighted average cost \$24 (private \$18 and public \$6) |
| Indonesia, 1998 [41] | Healthcare provider and Societal | Modeled national burden based on incidence 358–810/100 000 (200 M population)<br>**(All ages)** | **Direct costs:** Medication, surgery, bed charges, consultation<br>**Indirect costs:** Loss of work for patient/caregiver | **Mean outpatient cost estimated \$100-300**<br>**Mean inpatient cost w/o complication estimated \$200-1000**<br>**Mean inpatient cost with complication estimated >\$490** |
| India, 2024 [42] | Societal | 1 650 hospitalized fever cases<br>**(>6 months)** | **Direct Medical costs:** Medication, laboratory tests, bed charges<br>**Direct Non-medical costs**<br>**Indirect costs** | **Mean Out of pocket expense (OOPE)**<br>**S. Paratyphi infection \$124**<br>**S. Typhi infection \$105**<br>**Hospitalization of adult \$120**<br>**Hospitalization of paediatric \$79**<br>**Duration of hospitalization <3 days \$76, 3-5days \$115, >7days \$175**<br>**Patients that recovered without complications \$106** |
| | | | | **Patients with adverse outcomes (Sequalae/death/referred)**<br>\$139 |
| Tanzania, 2014 [43] | Societal | 17 blood culture–confirmed typhoid episodes at three Pemba district hospitals<br>(>2 months) | **Direct cost:**<br>Treatment expenditure<br>Travel expenditure<br>Meal's expenditure (before, during, and after hospital care)<br>**Indirect cost:**<br>Productivity loss (patient's absence from work)<br>Productivity loss (caregiver's absence from work) | **Total direct costs** \$38.11<br>**Total indirect costs**<br>**Children (<15 years):** \$216; **Adult:** \$162<br>**Total cost of illness**<br>**Children (<15 years):** \$265; **Adults:** \$193 |

### Cost of vaccine delivery/cost of vaccination of TCV

We identified four cost of vaccination studies [44–47] and one cost of vaccine delivery study [48], which are summarised in Table 6. Four of the five studies reported vaccine coverage based on primary data, with coverage ranging from 46.5% to 76% in India [44,47], 77% in Malawi [48], and 66.7% to 97.1% in Zimbabwe [46]. In contrast, Debellut et al. [45] used estimates for vaccine coverage rates ranging from 80% to 95%, depending on strategy and year.

**Table 6:** Summary of Cost of vaccination with TCV studies.

| Country, Year (Reference) | Strategies | Vaccination parameters | Source of costs | Key Findings (USD, 2024) |
| --- | --- | --- | --- | --- |
| India, 2020 [44] | **Campaign:** 9 months to 14 years | **Vaccine related costs:**<br>Cost of vaccine<br>Cost of Syringe<br>Cost of Safety Box<br>Vaccine wastage: 3%<br>Vial size: 5 doses<br>Vaccine coverage = 46.5%-76.1%<br>**Vaccination activities:**<br>Planning & preparation<br>Sensitization<br>Training<br>Microplanning<br>Social mobilization<br>Service delivery<br>Adverse event following immunization (AEFI) management.<br>Supervision<br>Monitoring | Navi Mumbai Municipal Corporation officials, the National Technical Advisory Group on immunization and other key stakeholders | **Total weighted average delivery cost per dose (including vaccine, syringes, and safety boxes):** \$5.40 (financial) and \$9.42 (economic).<br>**Total cost excluding vaccine, syringes, and safety boxes:** \$0.56 (financial) and \$1.78 (economic). |
| Malawi, 2022 [45] | **Routine:** >9 months<br>**Campaign:** >9 months to <15years | **Vaccine related costs:**<br>Cost of vaccine<br>Cost of Syringe<br>Cost of Safety Box<br>Vaccine & syringe wastage: 10%<br>Safety box wastage: 5%<br>Vial size: 5 doses<br>Vaccine procurement additional costs (% of vaccine price) 21%<br>Supplies procurement additional costs (% of supply price) 31%<br>**Estimated coverage:**<br>**Initial:** 95% in campaign<br>**Routine:** 2023, 80% 2024, 84% | Primary data collection, GAVI and UNICEF country office | **Cost Per dose:**<br>**Overall**<br>\$1.01 (Financial)<br>\$3.52 (Economic)<br>**Routine**<br>\$1.07 (Financial)<br>\$3.28 (Economic)<br>**Campaign**<br>\$1.00 (Financial)<br>\$3.55 (Campaign)<br>**Excluding the vaccine cost per dose:**<br>\$0.53 (Financial)<br>\$1.11 (Economic) |
| Zimbabwe, 2022 [46] | **Campaign:** >6 months to <45 years | **Vaccine related costs:**<br>Cost of vaccine<br>Cost of Supplies<br>Vaccine wastage: <0.01%<br>Vaccine coverage = 66.7%-97.1%<br>**Vaccination activities:**<br>Planning & preparation<br>Sensitization<br>Training<br>Microplanning<br>Social mobilization<br>Service delivery<br>Adverse event following immunization (AEFI) management.<br>Supervision<br>Monitoring | Surveillance data, WHO, UNICEF, Ministry of Health and Child Care, Harare City Health Department, | **Financial costs:**<br>**Mean cost per dose (vaccine and supplies)** \$2.99<br>**Mean cost per dose Supplies only** \$0.99 |
| India, 2023 [47] | **Campaign:** >9 months - <15 years | **Vaccines costs:**<br>Vaccine cost<br>Syringe cost<br>Safety box cost<br>Vaccine coverage achieved 71.0%<br>Vaccine wastage 3.1%<br>Syringe wastage 4.5%<br>**Activities:**<br>Planning and Preparation<br>Microplanning<br>Training<br>Sensitization<br>Social Mobilization<br>Service Delivery<br>Supervision and Monitoring<br>AEFI Preparedness and Management | Navi Mumbai Municipal Corporation officials, the National Technical Advisory Group on immunization and other key stakeholders | **Financial costs:**<br>**Weighted average delivery cost** \$0.48<br>**Total cost (vaccine+ delivery)** \$2.80-\$3.01<br>**Economic costs**<br>**Weighted average delivery cost** \$1.87<br>**Total cost (vaccine+ delivery)** \$5.43 - \$8.69 |
| Malawi, 2022<br>[48] | **Routine:** >9 months<br>**Campaign:** >9 months to <15years | **Program activities financial costs:**<br>Estimating demand for routine (Meetings)<br>Training (Fuel, maintenance & energy)<br>Management (Transport)<br>Social Mobilization (Stationary & communication)<br>Vaccine distribution & storage (Per diem)<br>Vaccine service delivery (Supplies)<br>**Program activities opportunity costs:**<br>Estimating demand for routine (Human resource)<br>Training (capital costs of vehicles & equipment)<br>**Others**<br>Supervision<br>Waste management<br>AEFI management (Campaign)<br>Record keeping & Monitoring<br>Vaccine coverage = 77% | Primary data collection using questionnaires from health facilities, EPI offices and vaccine stores. Secondary data used from different sources | **Costs excluding vaccine and vaccine supplies**<br>**Mean financial cost per dose:**<br>Health facility \$0.43<br>District level \$0.09<br>Zone level \$0.00<br>National level \$0.01<br>All levels \$0.53<br>**Mean economic cost per dose:**<br>Health facility \$2.66<br>District level \$0.17<br>Zone level \$0.01<br>National level \$0.04<br>All levels \$2.88 |

Cost data was obtained through a combination of primary data collection and secondary sources, including local health departments, national immunization technical advisory groups, and international organizations such as WHO, the United Nations Children’s Fund (UNICEF), and GAVI. In India, studies [44,47] primarily relied on data from local health officials and technical advisory bodies. Studies in Malawi [45,48] obtained primary level data and information sourced from international organizations, while the study in Zimbabwe [46] utilized data from the Ministry of Health.

The four cost of vaccination studies [44–47] incorporated costs associated with program-level activities (planning and preparation, microplanning, training, sensitization, social mobilization, supervision and monitoring, AEFI preparedness and management), as well as costs directly attributed to the vaccine (vaccine cost, syringe cost, safety box cost). Whereas the one cost of vaccine delivery study from Malawi [48] focused only on the cost of delivering the vaccine and excluded the cost of vaccines, syringes, safety boxes, and supplements.

The cost per dose varied significantly across regions. India [44,55] reported the highest financial costs, ranging from USD 2.80 to USD 5.40 per dose, and economic costs from USD 5.43 and USD 9.42 per dose. Malawi [45,48] reported lower financial costs (USD 0.87 -1.01 per dose) and economic costs USD 3.03 - 3.55 per dose). Zimbabwe [46] reported a mean cost of USD 2.99 per dose.

In terms of costs related to vaccine delivery only, they were relatively consistent across countries, with financial costs ranging from USD 0.48 to USD 0.99 [44–48] and economic costs between USD 1.11 and USD 2.88 per dose [44,45,47,48]. Table 6 summarizes the cost of vaccination studies and vaccine delivery studies.

### Public health impact studies

Two studies [34,49], evaluated the public health impact of different typhoid vaccination strategies for TCV. Both studies [34,49] employed a dynamic transmission model and incorporated parameters for demographic (birth and death rates), disease (infection duration, immunity duration, carrier probability), and transmission dynamic parameters (basic reproductive number, proportion symptomatic, chronic carrier infectiousness), along with vaccine characteristics (initial efficacy, duration of protection, and waning immunity), and antimicrobial resistance dynamics (resistance acquisition rates, transmission risks, and recovery rates). However, neither incorporated herd immunity as a model parameter.

Birger et al. [49] modeled a combination of routine vaccination at 9 months and campaign vaccination strategies targeting children aged 9 months to less than 15 years across 73 GAVI-eligible countries. In contrast, Kaufhold et al. focused on endemic countries, analyzing routine immunization at 9 months under varying coverage levels, from no vaccination to 100% coverage [34].

Higher coverage consistently led to reduced typhoid burden. Kaufhold et al [34], projected a 44% reduction in cases at 80% vaccination coverage and 58% reduction in cases at 100% coverage. A proportional decrease in AMR cases was also observed. Similarly, Birger et al.’s estimated a 46-74% reduction in typhoid cases,16.1% decline in resistant case proportions, 826,000 deaths averted, and 44.4 million DALYs averted [49].

Kaufhold et al [34] also highlighted that transmission rates, symptomatic proportions among vaccinated individuals, and chronic carrier infectiousness were the most influential parameters as per the sensitivity analysis. A summary of the findings from the public health impact studies is presented in Table 7.

**Table 7:** Summary of public health impact (modelling) of TCV studies.

| Country, Year (Reference) | Strategy | Model Type | Model Parameters | Key Findings |
| --- | --- | --- | --- | --- |
| Afghanistan, Angola, Armenia, Azerbaijan, Bangladesh, Benin, Bhutan, Bolivia, Burkina Faso, Burundi, Cambodia, Cameroon, Central African Republic, Chad, Comoros, Congo Rep, Côte d'Ivoire, Cuba, DR Congo, Djibouti, Eritrea, Ethiopia, Gambia, Georgia, Ghana, Guinea, Guinea Bissau, Guyana, Haiti, Honduras, India, Indonesia, Kenya, Kiribati, DPR Korea, Kyrgyz Republic, Lao PDR, Lesotho, Liberia, Madagascar, Malawi, Mali, Mauritania, Moldova, Mongolia, Mozambique, Myanmar, Nepal, Nicaragua, Niger, Nigeria, Pakistan, Papua New Guinea, Rwanda, São Tomé e Príncipe, Senegal, Sierra Leone, Solomon Islands, Somalia, Sri Lanka, Republic of Sudan, South Sudan, Tajikistan, Tanzania, Timor Leste, Togo, Uganda, Ukraine, Uzbekistan, Vietnam, Yemen, Zambia and Zimbabwe, 2022 [49] | **Routine + Campaign:** >9 months to <15years<br>**Time Horizon:** 10 years | **Dynamic transmission model**<br>S1, S2, +I1, S, I1,R, I2,S, I2,R, IT, + V1, V2, + CS, CR, + R | **Demographic parameters**<br>Crude birth rate (per year) ($\mu$)<br>Crude death rate (per year) ($\mu$)<br>**Fixed disease parameters**<br>Mean duration of infectiousness ($1/\delta$) 4 weeks<br>Fraction infected who become carriers ($\theta_a$)<br>Duration of immunity ($1/\omega$) 104 weeks<br>**Other Parameters**<br>Basic reproductive number 1.5, 2.5, 3.5, 7, 10.5,<br>Proportion symptomatic 0.01, 0.05, 0.10, 0.5,<br>Vaccination coverage 70–79%, 62–80%, 86–95%.<br>Relative infectiousness of chronic carriers ($r$)<br>**Vaccine-related parameters**<br>Initial efficacy of TCV ($v$) 87.5%, Uniform(80%, 95%)<br>Waning of vaccine-induced immunity ($\omega V$) 0.0672 per year, Gamma (1.40, 0.0479)<br>**Resistance parameters**<br>Rate of resistance acquisition from treatment 0.01–5 per week,<br>Relative risk of transmission for resistant strain 0.3–3,<br>Rate of recovery from primary infection with a resistant strain 0.075–0.7 per week,<br>Rate of recovery from subclinical infection with a resistant strain 0.075–0.7 per week. | **General**<br>- Predicted cases averted 66.7 million (48.1–88.3 million) cases i.e. 46–74%.<br>- Predicted deaths averted 826000 (381000–2.4 million) deaths.<br>- Predicted DALYs averted 44.4 million (18.7–143 million) DALYs.<br>**Anti-microbial resistant typhoid**<br>- Fluoroquinolone non-susceptibility (FQNS) cases averted 42.5 million (24.8–62.9 million) cases.<br>- Multidrug resistance cases averted 21.2 million (16.4–26.5 million) cases<br>- Overall antimicrobial-resistant typhoid fever averted 53.5 million (37.3–75.3 million) cases<br>- Predicted reduction of resistant cases proportion 16.1% (0–49%)<br>- Prevalence of resistant cases expected to decrease by 1.1% for every one-unit increase of $R_0$. |
| Endemic areas, 2019 [34] | **Routine** only at 9 months<br>**Time Horizon:** 10 years<br>**Scenarios:** no vaccination, 30% coverage, 50% coverage, 80% coverage, and 100% coverage | **Dynamic transmission model**<br>S1, S2, +I1, S, I1,R, I2,S, I2,R, IT + V + CS, CR, + R | **Demographic parameters:**<br>Birth and death rate<br>**Fixed disease parameters:**<br>Duration of infectiousness for first antimicrobial-sensitive inf.: 4 weeks<br>Fraction who experience disease-induced mortality<br>Duration of temporary immunity following recovery from natural infection, 104 weeks<br>**Other parameters:**<br>Transmission parameter 0.3 (0.05–0.5)<br>Vaccine efficacy (initial) 0.95 (0.8–1)<br>Reduction in risk of infection for vaccinated 1 (0.5–1)<br>Duration of protection from vaccination 19.2 years<br>Fraction symptomatic 0.2 (0.1–1)<br>Fraction symptomatic for vaccinated individuals 0.05<br>Fraction treated 0.75 (0–1)<br>Duration of infectiousness with treatment 1 week<br>Relative infectiousness of chronic carriers 0.35<br>Fraction that become carriers from 1st, antimicrobial-sensitive infection 0.03<br>Relative rate of recovery from second infection 1<br>**Resistance parameters**<br>Rate of treatment-induced/acquired resistance<br>Natural recovery rate from first, antimicrobial-resistant infection 0.2 (0.1–0.5)/ week<br>Relative infectiousness of resistant strain(s) 0.9<br>Fraction becomes carriers from the 1st, AMR infection | **No Vaccination Group:** 483 cases per 100,000 person-years.<br>**80% Coverage:** Reduced to 270 cases per 100,000 person-years, a 44% decrease.<br>**100% Coverage:** Further reduced to 202 cases per 100,000 person-years, representing the maximum reduction of 58%.<br>The most impactful parameters on the percent of cases averted were the **transmission rate** and **proportion of cases among vaccinated individuals that are symptomatic**, followed by the **duration of immunity** afforded by vaccination, and the **relative infectiousness of chronic carriers**. |

### Economic evaluations (cost-effectiveness analysis)

There were nine cost-effectiveness analyses [20,27,29,35,50–54] included in this review. The studies employed models incorporating a range of demographic, disease transmission, and vaccine-related parameters. Sensitivity analyses were conducted by all the studies to identify key drivers influencing the cost-effectiveness related outcomes.

The vaccination strategies were compared to a baseline “do-nothing” scenario to estimate the incremental benefits of TCV. Five studies assessed routine vaccination targeting children aged 6 months to 1 year, in combination with campaign-based approaches targeting broader age groups (such as 9 months to 5 years or up to 30 years) [20,29,35,50,54]. Three studies evaluated campaign strategies delivered through school- or community-based approaches [51–53], while Chauhan et al. varied vaccine efficacy within routine-only approaches to simulate different vaccine strategies [27]. Health outcomes were primarily measured in DALYs [20,29,35,50,51,53,54], with two studies opting to use quality-adjusted life years (QALYs) [27,52].

Case reductions ranged from 2% (routine-only, pessimistic scenarios) to 94% (routine plus campaigns) [20,27,29,35,51,53,54]. Deaths averted ranged from 0% to 36% for routine-only vaccination in low-incidence areas, to 17% to 100% when combined with campaigns [20,27,29].

In terms of cost-effectiveness, the reported ICER varied based on typhoid vaccination strategy used. Antillón et al. [29] and Chauhan et al. [27] reported all strategies to be cost-effective, while Lo et al. [35] and Burrows et al. [54] found routine-only vaccination as most cost-effective. In contrast, four studies [20,50,51,53] concluded that combining routine vaccination with campaigns was more cost-effective, with routine-only strategies often dominated. Amongst these four studies, Weyant et al. [53] used targeted campaigns that included schools, and Ryckman et al. [51] included both community and school-based campaigns. The study done in the Lao PDR [52] was the only study that showed no vaccination as the most cost-effective strategy, primarily due to the low incidence of typhoid in the country.

Studies adopting a societal perspective [27,35,51,53,54] consistently reported lower ICERs than those using a healthcare provider perspective. This was highlighted by Chauhan et al., [27] reporting ICER values ranging from $789 to $2,642 per QALY (provider perspective) versus -$1,218 to -$4,251 per QALY (societal perspective) [27]. Similarly, Ryckman et al. [51] reported ICERs of USD 1,142 per DALY averted (societal) compared to USD 2,228 per DALY averted (payer), and Weyant et al. found a societal ICER lower than the payer estimate of USD 567 per DALY averted [51,53].

High-incidence areas consistently demonstrated greater cost-effectiveness for TCV compared to low-incidence areas. Notably, five studies [20,27,29,35,51] concluded that in high-incidence settings, TCV was not only cost-effective but also cost-saving, offering greater health benefits at lower costs than alternative strategies. These studies are summarized in Table 8.

**Table 8:**
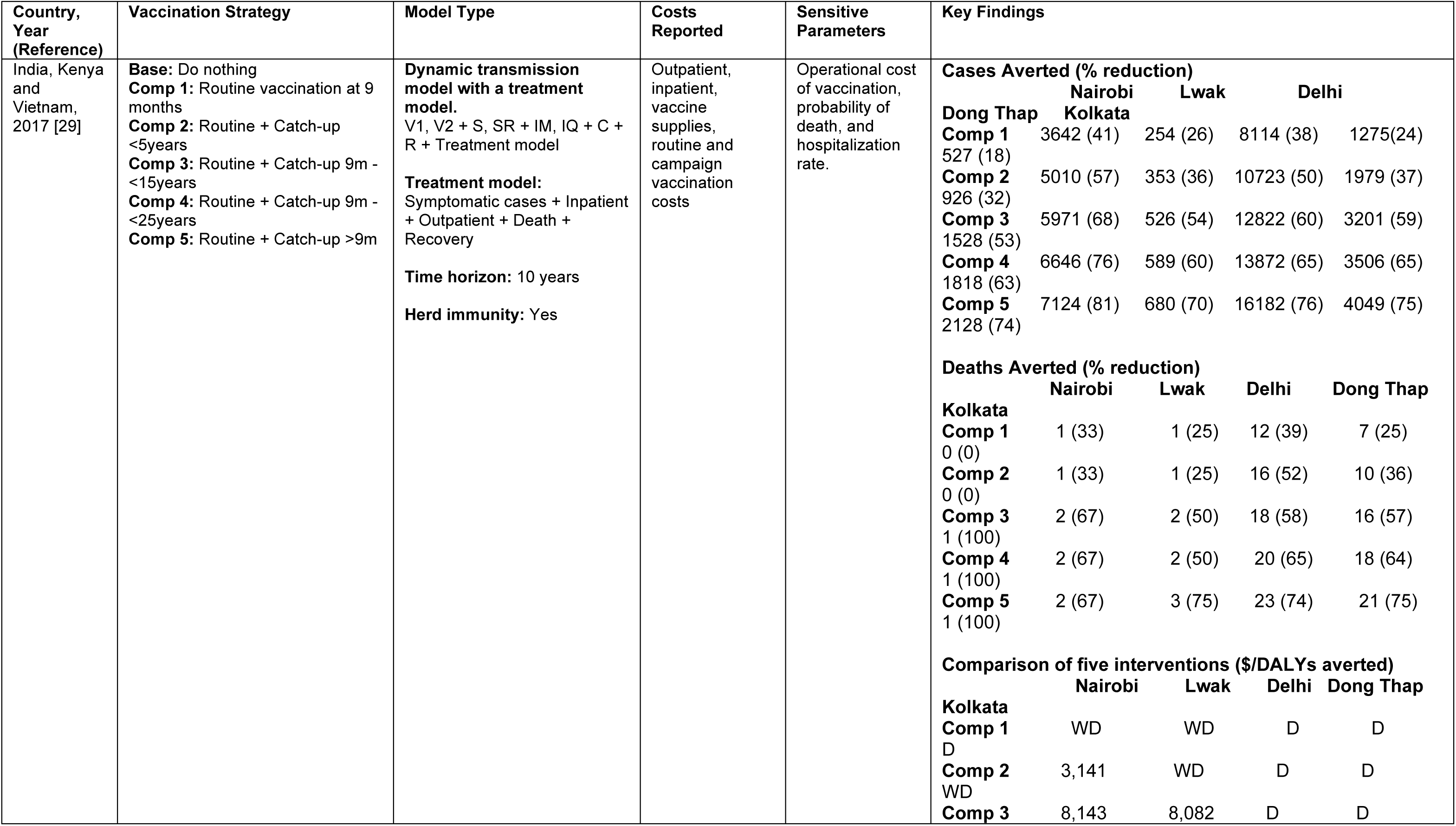
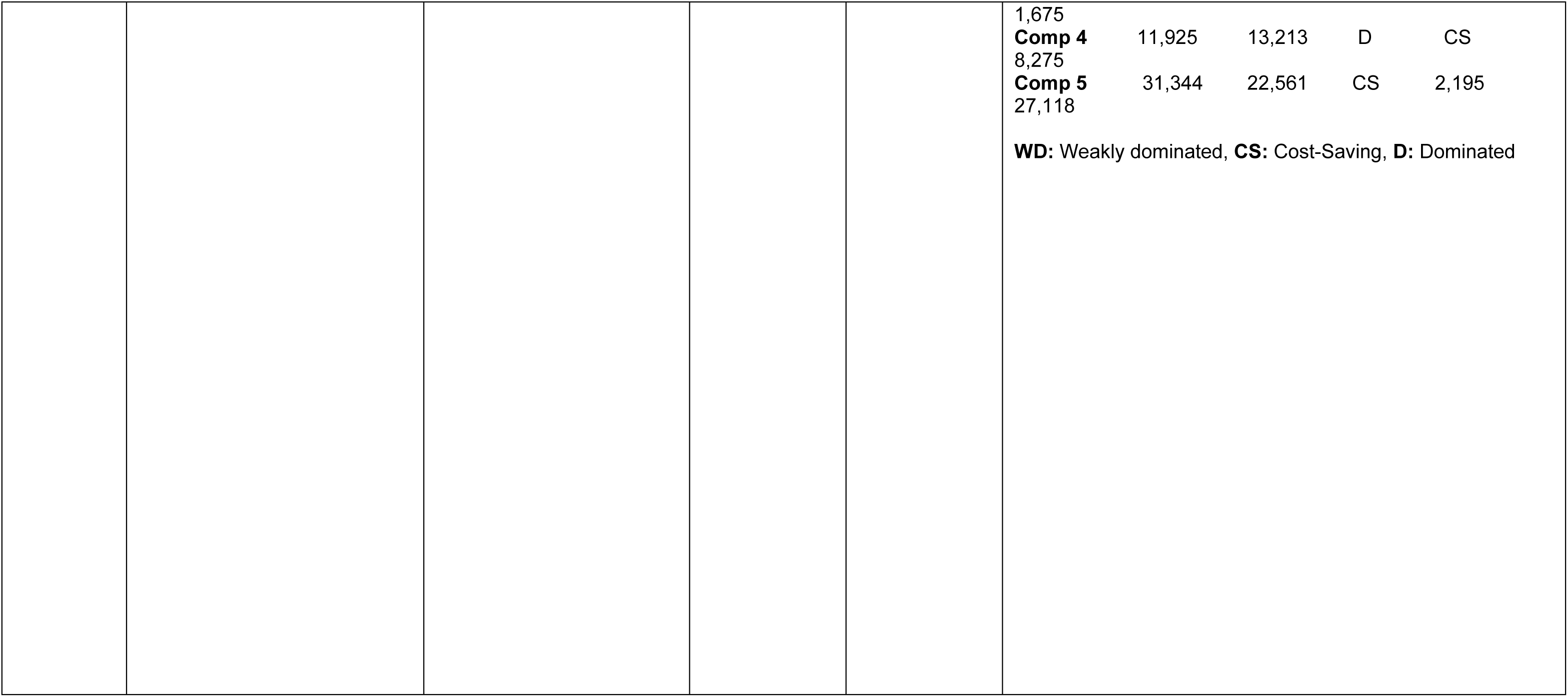
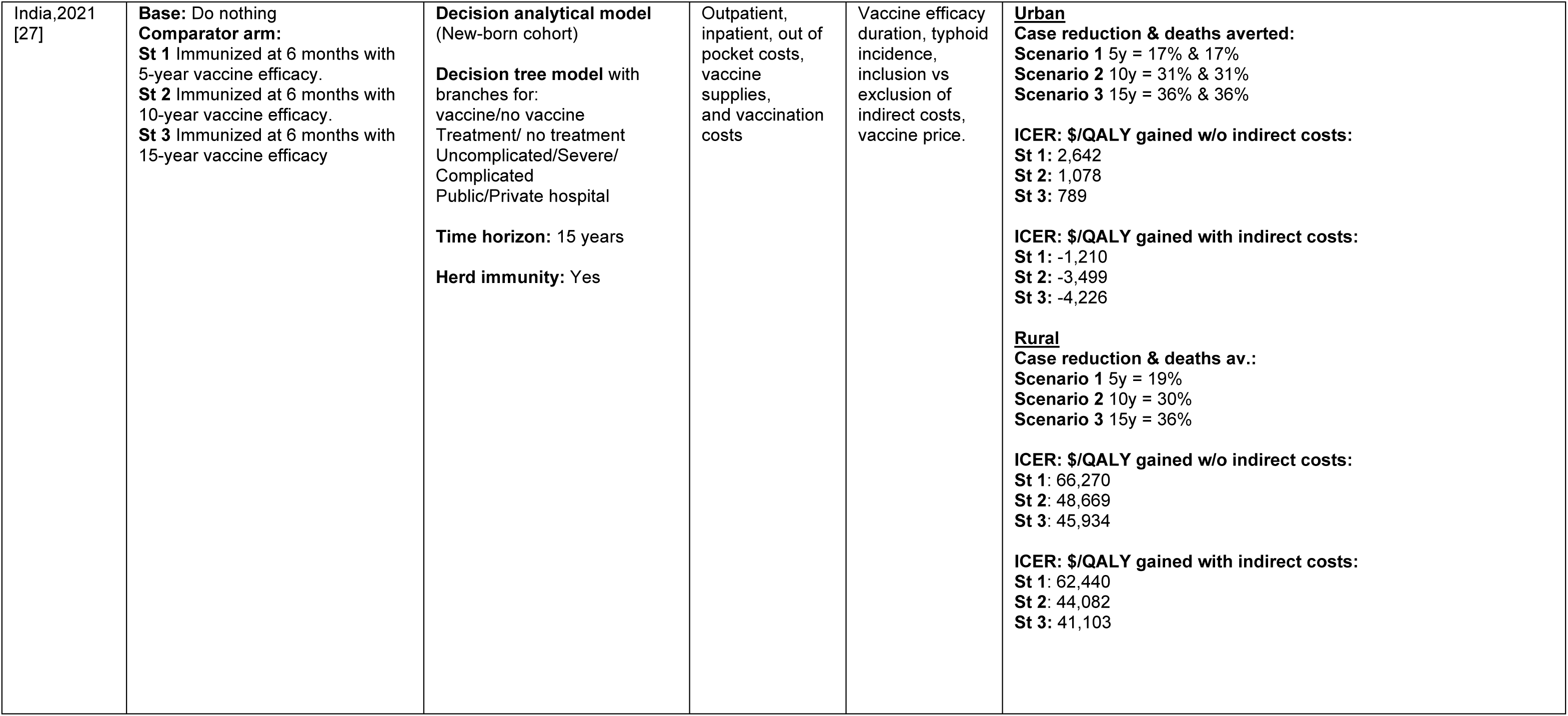
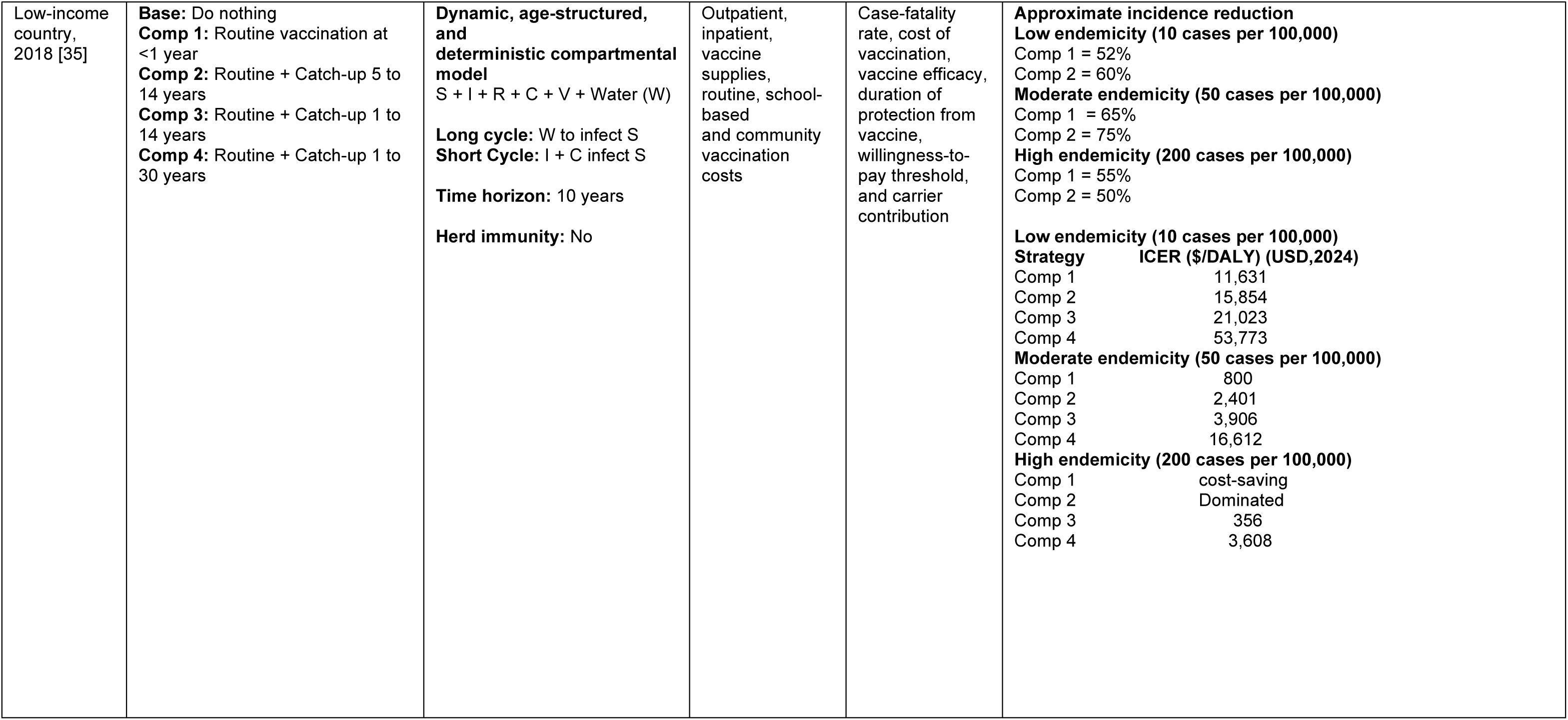
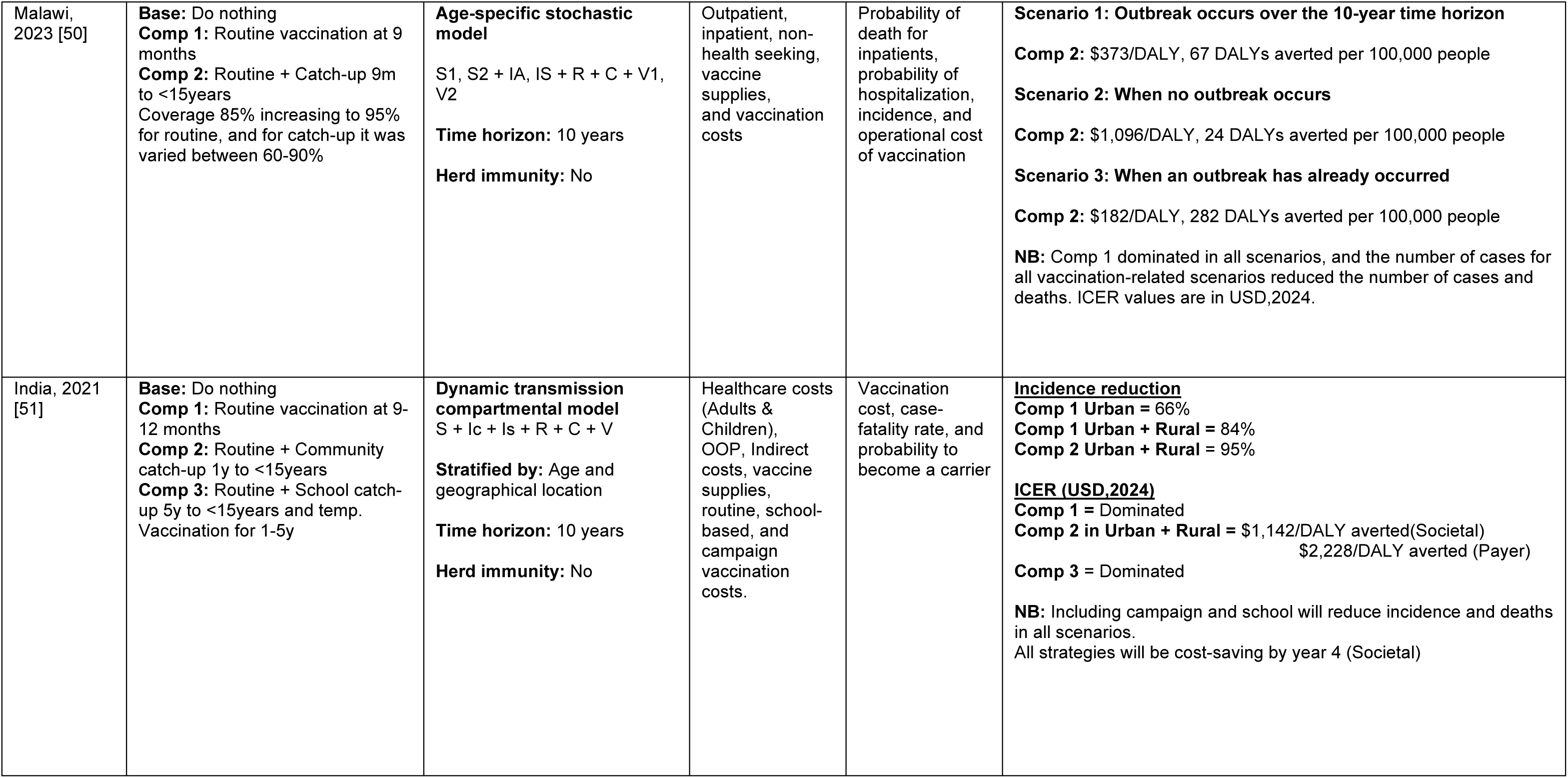
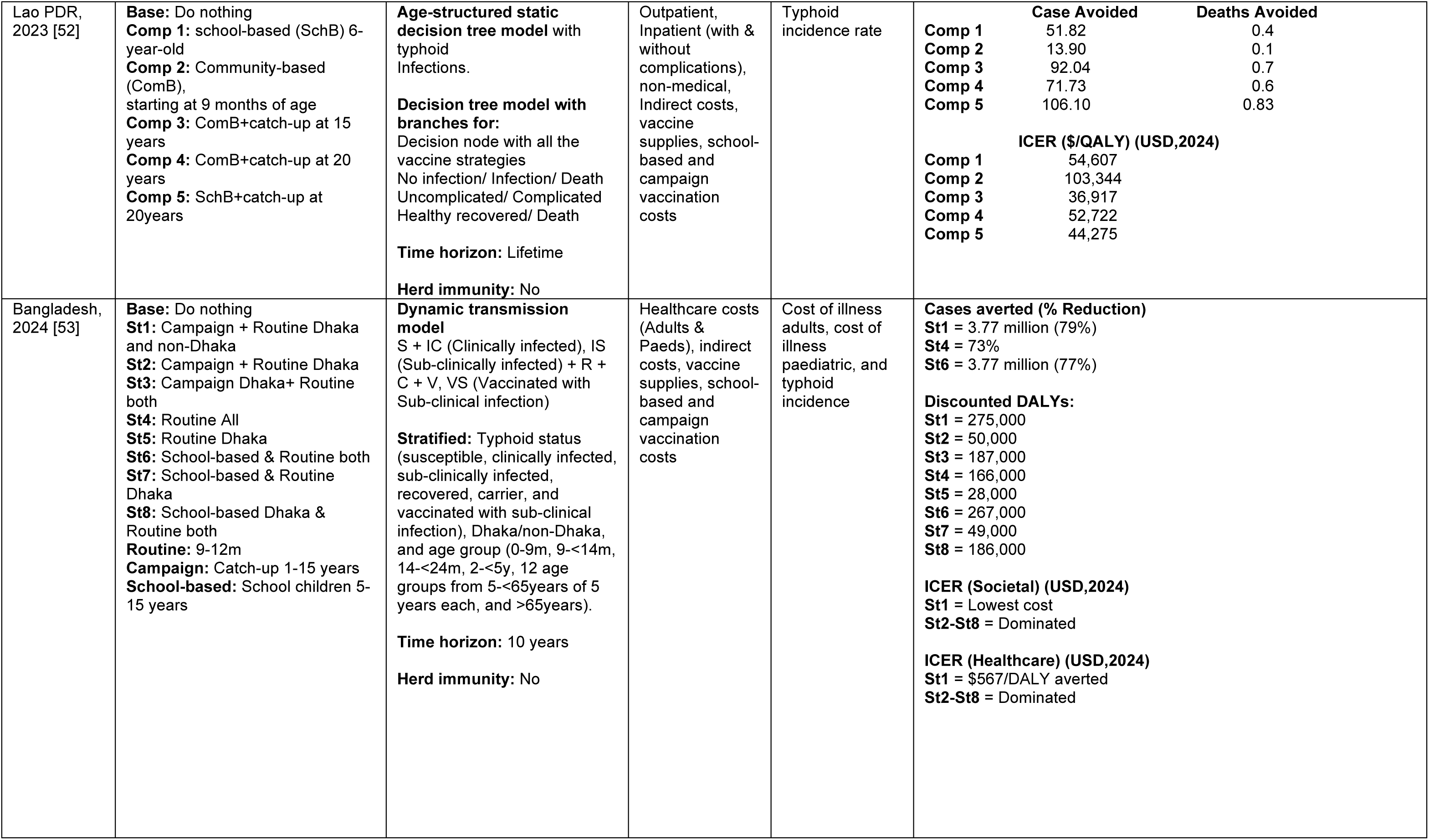
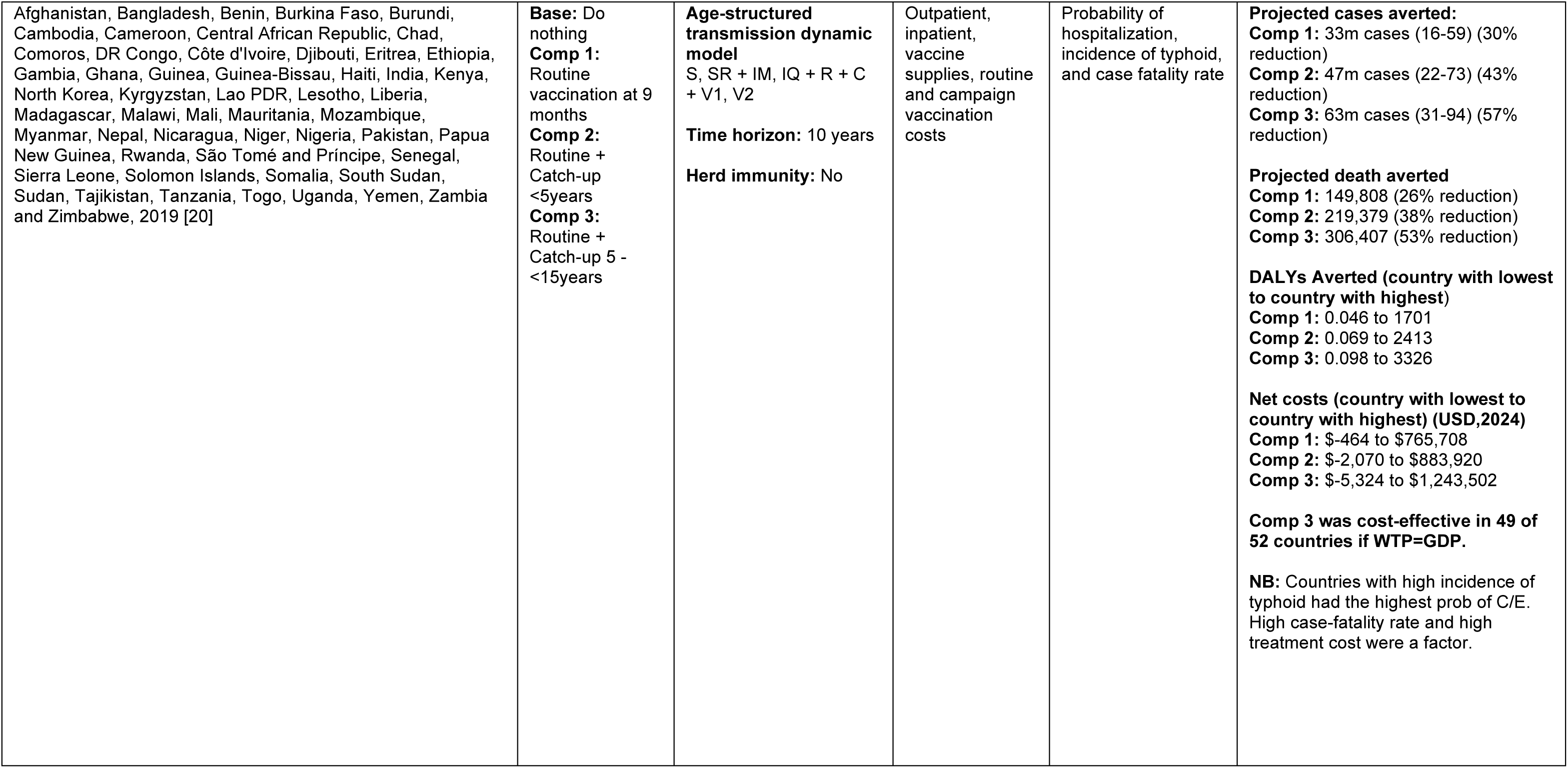
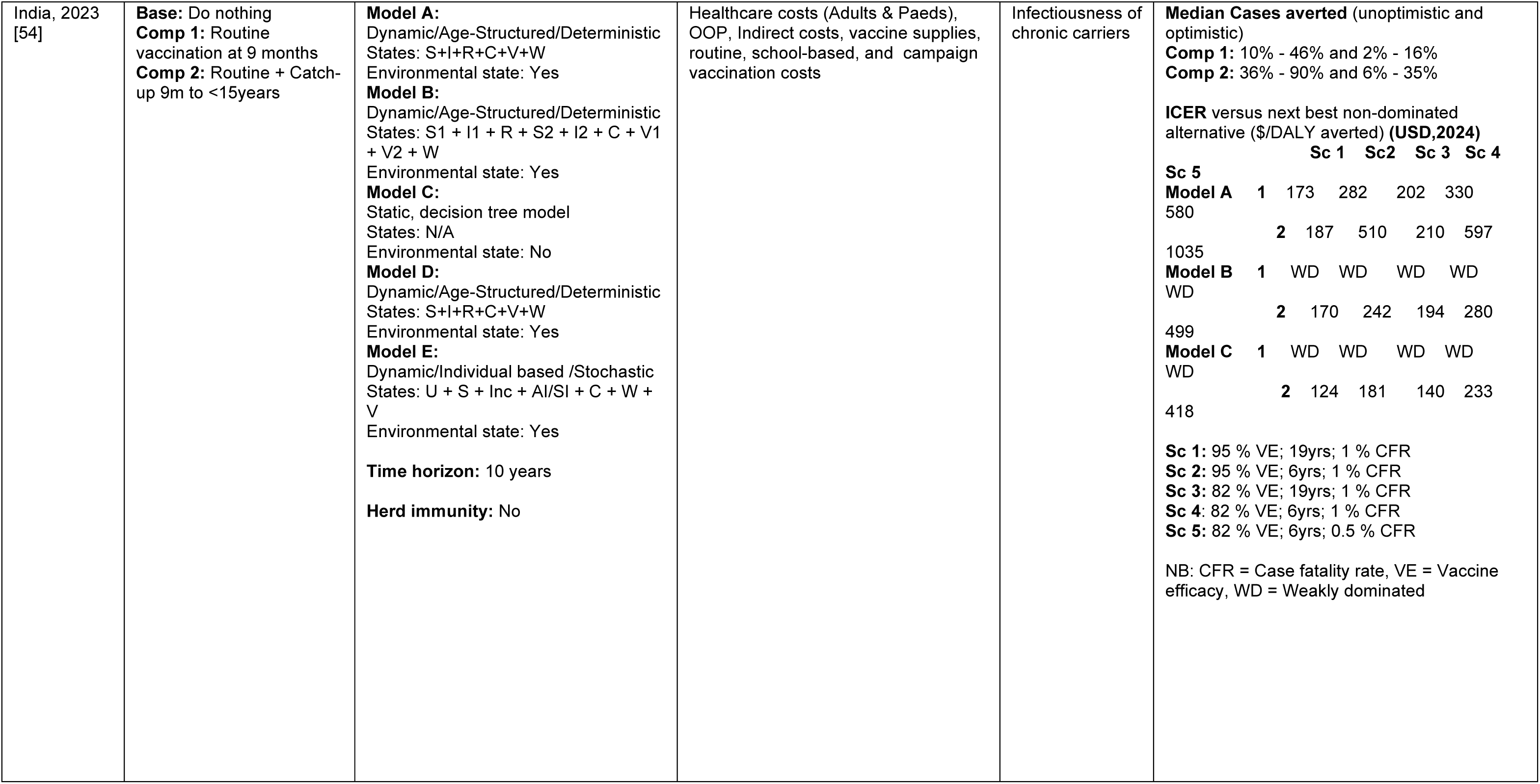
Summary of Economic Evaluation of TCV.

## Discussion

This scoping review systematically mapped existing literature on the economic and public health impact of TCV, highlighting key findings, gaps, and implications for future research and policy.

Of the 24 included studies, 19 were from South Asia and Sub-Saharan Africa [20,27,29,36–38,40,42–46,48–51,53–55]. This concentration can be attributed to the significant public health burden of typhoid in these regions and the ongoing vaccination-related initiatives [2,3]. However, regions such as Latin America and parts of Asia, where typhoid is endemic, were notably underrepresented. For evidence-based decision-making in these regions, more studies are needed in the local context.

Studies focusing on the COI of typhoid [36–43] highlighted the substantial economic burden of typhoid fever. Higher costs were generally associated with inpatient care compared to outpatient care, and indirect costs often exceeded direct medical costs [37–39,43]. Of the five studies that reported costs for both children and adults [36–38,42,43], only one reported the cost to be higher in children, which was attributed to longer hospitalization, higher medical and indirect costs [43]. The remaining four studies reported higher costs in adults [36–38,42], with three reporting higher direct medical costs [36–38], and two studies reporting higher indirect costs [38,42] in adults when compared to children. Studies adopting a societal perspective further emphasized the burden, reporting that direct non-medical and indirect costs comprised 34-98% of total treatment costs [36–39,43]. Beyond the use of a societal perspective, several other factors contributed to the wide variation in cost estimates across studies. For instance, three studies included the cost of surgical interventions, which significantly increased overall treatment costs [36,38,41], whereas 4 studies included costs from private or higher-tier hospitals [36,38,40,41], where the cost of treatment tends to be higher than in public healthcare facilities. The cost variations could also be attributed to regional differences in the prices of goods and services, as these costs are context-specific. This review underscores the importance of having local COI data to help understand the burden households face. For example, one study in India found that households paid as much as USD 175 out of pocket for typhoid treatment [42], a cost that is beyond the reach of most families [56]. Policymakers can draw on COI data to design financial protection measures and shape equitable, effective health policies.

Studies on the cost of vaccination [44–47] and vaccine delivery [48] highlighted different factors influencing implementation costs across various settings. Variations in the cost of delivering TCV were associated with the vaccination strategy used (routine or campaign-based), logistical challenges, vaccine wastage rates, human resource costs, and regional differences in vaccine pricing. The higher costs reported in India were mainly attributed to the cost of the vaccine and supplies (USD 3.69 compared to an estimated USD 2.40 in Malawi [45]), low vaccine coverage in high end neighborhoods due vaccine hesitancy and extra personnel needed for mobilization, with the cost of delivery rising to USD 4.90 (economic cost) per dose in these neighborhoods [47]. In contrast, Malawi and Zimbabwe reported lower costs due to higher coverage estimates and using staff from the existing immunization program [45]. Vaccine delivery costs varied based on specific activities, such as community outreach, cold chain maintenance, adverse event monitoring, and healthcare worker training, as well as broader systemic factors including population characteristics, vaccine hesitancy, and healthcare capacity. As countries transition out of GAVI support [57], future cost-effectiveness analyses should focus on these cost drivers using local data to better inform policymakers.

Dynamic models were used in the two public health impact studies [34,49] and five economic evaluations [20,29,35,51,53,54] to simulate changes in typhoid transmission, population structure, and immunity levels over time. Dynamic models are preferred over static models because they effectively capture the time-dependent changes associated with typhoid transmission and the indirect effects of vaccination [58]. All models were a variation of the Susceptible-Infectious-Recovered (SIR) model [59], with additional states such as Vaccination (V), Chronic Carrier (C), and Infection through water sources (W) [20,34,35,49,50,50,51,53,54] being included. However, due to limited data availability, the models [34,49] relied on assumptions about typhoid transmission rates, antimicrobial resistance, health-seeking behaviour, vaccine coverage, and vaccine efficacy. The potential impact of vaccine hesitancy and the effect of herd immunity were frequently excluded, limiting the models’ real-world applicability. To address this, countries can strengthen surveillance systems, research the level of vaccine hesitancy, and conduct studies to understand vaccine efficacy in the local population.

The nine economic evaluations [20,27,29,35,50–54] identified in this scoping review reflect the growing interest in assessing the value for money of TCV, especially given its recent introduction and adoption. Before the WHO’s 2017 recommendation to include TCV in routine immunization programs [21], earlier types of typhoid vaccines were not widely used due to limitations in effectiveness, age restrictions, and multiple doses [11,12]. Despite the novelty of TCV, the emerging work on economic evaluations suggests strong interest from researchers and policymakers, underscoring a recognition of TCV’s potential to yield substantial public health and economic benefits.

Variations in the reported ICERs across the economic evaluations were primarily driven by the vaccine strategy employed and the incidence of typhoid. TCV was shown to be more cost-effective in high-incidence and urban settings, where the disease burden is greater, population density is higher, and sanitation infrastructure is often inadequate. These characteristics of urban settings offer a greater opportunity to avert cases and death, making these areas a key target for cost-effective vaccination strategies [60,61]. Four studies reported TCV to be cost-saving in these urban areas, as the reduction in treatment costs outweighed the costs associated with vaccination [27,29,35,51]. This further enforces the need for policymakers to consider vaccine strategies that target densely populated, urban areas.

In terms of vaccine delivery strategy, the review showed that combining routine vaccination alongside catch-up campaigns targeting broader age groups was generally more cost-effective than routine vaccination alone [29,50,51,53]. However, some studies identified routine vaccination as the most cost-effective strategy in specific scenarios [27,35,52,54]. These findings underscore the need for policymakers to utilize context-specific vaccine strategies to maximize both health outcomes and economic impact.

A subset of studies that reported ICERs from both societal and healthcare provider perspectives showed that the ICERs were significantly lower when the societal perspective was used, primarily due to the inclusion of indirect costs, which were substantial [27,51,53]. Researchers should adopt the societal perspective in future CEAs of TCV to fully capture the economic benefits of vaccination.

Sensitivity analyses identified several key parameters that the ICER was sensitive to. These included typhoid incidence rates, vaccine efficacy, coverage levels, case-fatality rates, and treatment costs. Higher incidence rates improved cost-effectiveness by increasing the number of cases averted [27]. While greater vaccine efficacy and higher coverage enhanced the health gains from vaccination [62]. High case-fatality rates and high treatment costs also improved the cost-effectiveness of vaccination by highlighting the vaccine’s potential to prevent deaths and reduce health-related expenditures.

A notable finding of this review was the consistent underrepresentation of herd immunity, health-seeking behavior, and unreported deaths outside a hospital setting in the included models. These omissions could have led to an underestimation of the broader public health and economic benefits of TCV, as reducing disease transmission through vaccination confers indirect protection to unvaccinated individuals [63]. Similarly, the impact of TCV on antimicrobial resistance (AMR) was insufficiently explored in the cost-effectiveness studies, with only the two modeling studies explicitly considering this parameter [34,49]. Given the rising prevalence of multidrug-resistant *S. Typhi* and the associated treatment challenges [64], incorporating AMR into future models would be essential for accurately capturing the full economic value of TCV.

Several critical areas for future research have been highlighted in this review. First, the use of a lifetime horizon should be explored as it will highlight the long-term economic and public health benefits of TCV, which is essential to inform sustainable immunization policies. Second, there is a need for models to integrate parameters such as herd immunity, AMR, and account for regional variations of typhoid. Third, more primary-level cost data for vaccine delivery and program implementation in diverse settings are required to refine cost-effectiveness analyses. Lastly, evaluating the equity implications of TCV introduction, particularly in low-resource settings, may provide critical insights to guide interventions that reduce health disparities and support equitable global health decision-making.

This scoping review had some limitations. Although we did not apply language restrictions in our database search, we only included articles published in English, so studies reported in other languages may have been missed. We did not use formal quality-appraisal tools for cost-of-illness, cost-of-vaccination, or public-health-impact studies, and therefore could not evaluate the methodological rigor of individual analyses. We also did not search the grey literature, which may have led to the omission of unpublished program evaluations or reports.

## Conclusion

This scoping review highlights the role of vaccination in reducing the burden of typhoid and the regional/inter-regional differences in the cost-effectiveness of the vaccine, emphasizing the importance of using local data. Literature shows that TCV can not only be cost-effective but also cost-saving in high-burden settings. To maximize the impact of TCV, policymakers should prioritize its implementation in high-burden areas and employ targeted vaccination strategies to achieve this goal. Future research should focus on developing comprehensive models that capture the broader economic and public health impacts of TCV and incorporate subnational data to ensure evidence-based decision-making for sustainable immunization programs.

## Data Availability

All data extracted and analyzed during this scoping review are included in the article and its supplementary materials.

## Supporting Information Captions

**S1 Appendix. Detailed search strategy**

Search strategy used across PubMed, Web of Science, HTA, Scopus, and NHS EED databases to identify studies.

**S2 Appendix. PRISMA-ScR checklist**

Completed PRISMA-ScR checklist showing compliance with scoping review reporting guidelines, with corresponding manuscript page numbers.

**S3 Appendix. Microsoft Excel template for data extraction**

Data extraction form to capture study ID, vaccine strategy, model type, model parameters, cost of illness, cost of vaccination, and key findings.

**S4 Appendix. Drummond checklist for assessing cost-effectiveness**

Checklist used to assess the methodological quality of economic evaluations, rated on a scale of 1–10.

**S5 Table. Master data extraction table (Excel file) <u>Comprehensive Data Extraction Table (Excel file)</u>**

Table that consolidates all extracted study data, including unadjusted cost values, before outcome grouping.

